# Reading Between the Signs: Predicting Future Suicidal Ideation from Adolescent Social Media Texts

**DOI:** 10.1101/2025.08.28.25334643

**Authors:** Paul Blum, Enrico Liscio, Ruixuan Zhang, Caroline Figueroa, Pradeep K. Murukannaiah

## Abstract

Suicide is a leading cause of death among adolescents (aged 12–18), yet predicting it remains a significant challenge. Many cases go undetected because young people often do not contact mental health services. In contrast, young people often share their thoughts and struggles online in real time. To utilize this communication channel, we propose a novel task and method: predicting suicidal ideation and behavior (SIB) from online forums *before* an adolescent explicitly expresses suicidal ideation on a forum. This predictive framing, where self-disclosure is not used as input at any stage, is largely unexplored in the suicide prediction literature. We introduce Early-SIB, a transformer-based model that sequentially processes the posts a user writes and engages with to predict whether they will write a SIB post. Our model achieves a balanced accuracy of 0.73 in predicting future SIB on a Dutch youth forum, demonstrating that such tools can offer a meaningful addition to traditional methods.

## 1 Introduction

Suicide is a pressing issue, globally being the third leading cause of death among 15–29 year olds (World Health Organization, 2024). Moreover, each suicide is estimated to meaningfully affect 135 individuals around the victim (Cerel et al., 2019). However, predicting suicide is a significant public health challenge. 60% of people who die by suicide have not expressed suicidal ideation as documented in their medical records or inquired through a semi-structured interview (McHugh et al., 2019). Furthermore, our ability to predict suicide has not improved over the past 50 years (Franklin et al., 2017). Thus, there is an urgent need for innovative methods to detect suicide risk in adolescents.

Research on suicide has mainly focused on assessing a person’s risk of suicide using the so-called “risk factors” (Turecki et al., 2019). These are traits or conditions thought to increase the likelihood of suicidal behavior. These factors have informed widely adopted guidelines (American Association of Suicidology, 2023; World Health Organization, 2024; American Foundation for Suicide Prevention, 2025; National Institute of Mental Health (NIMH), 2025). For adolescents, risk factors associated with suicide include sex, depression, anxiety, adverse childhood experiences, self-hate, and self-injury (Cheng et al., 2025). However, a meta-analysis of decades of suicide research reveals that risk factor-based prediction performs only slightly better than chance (Franklin et al., 2017). The guidelines often lack specificity, classifying nearly anyone with mental illness, chronic physical illness, life stress, special population status, or access to lethal means as being at risk (Franklin et al., 2017). A more recent meta-analysis (Schafer et al., 2021) directly compares theory-driven approaches with machine learning (ML) models, and finds that the latter provides substantially superior predictive accuracy for suicide ideation, attempts, and death.

Given the widespread use of social media and increased self-disclosure during the years of adolescence (Vijayakumar and Pfeifer, 2020), social media data could prove to be a valuable source for identifying suicide risk in teenagers. Teenagers often do not disclose risk factors to their physicians; instead, many share them on social media (Pourmand et al., 2019), often making peers the first recipients of their distress calls (Belfort et al., 2012; Vijayakumar and Pfeifer, 2020). Additionally, over two-thirds of people who die by suicide have had no contact with professional mental health care in the year before their death (Stene-Larsen and Reneflot, 2019), essentially making them overlooked by traditional healthcare systems. Even for those who seek professional help, the time between healthcare interactions is immense compared to social media interactions (Coppersmith et al., 2018).

Suicidal Ideation and Behaviour (SIB) refers to thoughts, plans, or actions related to suicide (Posner et al., 2010). Ideation is one of the earliest and typically unobserved stages in the progression of an individual’s suicidal process (Wasserman et al., 2012). SIB, which also includes suicide attempts, is one of the strongest predictors of death by suicide (Klonsky et al., 2016). If we can predict SIB early, we could enable more timely intervention.

We address an under-explored challenge and propose a method to approach suicide prediction in adolescents: predicting future SIB based solely on a user’s social media post history. Unlike existing approaches, our method does not rely on any self-disclosure of SIB as input to the model, effectively moving the prediction one step earlier in the time-line. To perform this task, we introduce EARLY-SIB, a transformer-based model designed to predict SIB from the posts that a user has written and interacted with. We experiment with EARLY-SIB on De Kindertelefoon^1^, a Dutch adolescent help forum, and show that SIB can be predicted from a user’s post history. We then employ SHAP, an explainable AI technique, to reveal how the model consistently relies on a combination of the posts to make its predictions, rather than on a few significant posts, underscoring the complexity of the task. These findings suggest that ML tools applied to social media posts may complement traditional risk factor-based approaches for suicide prediction.

## 2 Related Works

We first review traditional clinical methods of identifying suicide risk before turning to recent work using natural language processing (NLP).

### 2.1 Clinical Methods

The U.S. Food and Drug Administration (2012) considers the Columbia Suicide Severity Rating Scale (C-SSRS) to be the gold standard for suicide risk assessment in clinical trials. C-SSRS provides a list of questions, e.g., “have you had any thoughts about killing yourself?”, to identify suicidal ideation and behavior (Posner et al., 2010). The European Psychiatric Association (Wasserman et al., 2012) recommends that suicide risk be assessed through clinical interviews that evaluate the patient’s psychological and social functioning. This includes assessments of personality, evaluations of socio-economic status, and the use of psychometric instruments such as scales (Hamilton, 1960; Paykel, 1976; Linehan et al., 1983; Patterson et al., 1983; Posner et al., 2010). There are also biological measurements, but they are only possible at specialized clinics and mainly used in research settings (Wasserman et al., 2012).

Most of these risk assessment tools rely on directly asking individuals about suicidal thoughts or evaluating predefined psychological factors. Instead of relying on individuals to disclose their mental state, our method infers risk from their online behavior and avoids dependence on predefined risk factors by leveraging ML to learn SIB patterns.

### 2.2 NLP for Suicide Detection

Recent years have seen growing interest in using NLP to assess suicide risk. Many studies use patients’ electronic medical records (Carson et al., 2019; Bittar et al., 2019; Zhong et al., 2019; Tsui et al., 2021; Singh Rawat and Yu, 2022; Zandbiglari et al., 2025) or chat messages between help-seeks and providers (Bialer et al., 2022; Broadbent et al., 2023; Qiu et al., 2024). However, these approaches neglect people who never seek help or have limited contact with healthcare professionals.

Another approach is using social media platforms like Twitter and Reddit, e.g., by classifying posts into containing suicidal thoughts or not (Cao et al., 2019; Mathur et al., 2020; Roy et al., 2020; Haque et al., 2022) or classifying users into different risk levels of suicide (Mohammadi et al., 2019; Sawhney et al., 2020, 2021a,b). Datasets have supported this direction by providing Reddit posts which have been expert-labeled into risk categories (Zirikly et al., 2019; Gaur et al., 2019; Park et al., 2020). While these approaches demonstrate the feasibility of detecting users at risk, they do not predict future disclosure of SIB. This limits their utility for early prevention or broader screening.

A smaller number of studies have attempted to infer suicide risk early. Coppersmith et al. (2018) and work in the CLPsych Task 2021 (MacAvaney et al., 2021; Gollapalli et al., 2021; Gamoran et al., 2021; Morales et al., 2021; Wang et al., 2021) used social media posts from users with actual suicide attempts. However, such data may already include explicit self-reports of suicide, limiting its usefulness for early detection. In contrast, we aim to predict self-reports before the user ever shares them, moving one step earlier in the timeline.

Recent efforts under the CLPsych Task 2024 (Chim et al., 2024) have been made to provide explanations of the model’s risk prediction. They highlight salient sub-phrases within suicidal Reddit posts and summarize that evidence at the user level (Koushik et al., 2024; Alhamed et al., 2024; Gyanendro Singh et al., 2024). In contrast, our work explains predictions in an early prediction setting, where we highlight earlier interactions that do not contain SIB but are indicative of future risk.

### 2.3 NLP for Mental Health Detection

Aside from SIB, NLP has been used for early detection of mental health issues, including depression (Zogan et al., 2022; Wang et al., 2024; Hadzic et al., 2024), eating disorders (López-Úbeda et al., 2021; Mármol Romero et al., 2024), and gambling addiction (Perrot et al., 2022; Smith et al., 2024), with one of the most important forums being ERisk@CLEF (Montejo-Ráez et al., 2024).

We draw on architectural ideas from Zhang et al. (2024), who proposed a model for detecting users with depression on social media. While they use posts directly indicative of depression as input, their architecture proves to be a valuable starting point for our task, because it is capable of taking multiple posts from a user’s history as input.

## 3 Dataset

We use data from the Kindertelefoon^1^, a Dutch forum intended as a space where adolescents (12–18 years of age) can share their feelings and experiences that they may find difficult or uncomfortable to discuss otherwise. Some of the most popular subforums on the platform include emotional problems and feelings, relationships and love, gender identity, and sexuality. Users can anonymously participate in the forum by creating a post or replying to an existing one. Each post includes a title and a body; it may contain tags (keywords) selected by the user to describe the content. Replies consist only of a body of text. In this work, we use the term *interaction* to refer to both posts and replies.

We first signed a collaboration agreement with the Kindertelefoon, including permission to scrape their forum (which is publicly available data), and obtained approval from the ethics committee of the home university of the leading author. We then extracted over 37,000 forum posts, along with all corresponding replies. All usernames were pseudonimized. We proceeded by labeling a small *post-level* dataset, which we use to train a model to detect SIB posts. This model is then used to create a *user-level* dataset that excludes any SIB posts. Data and annotations will be available under restricted access, as per agreement with the Kindertelefoon.

### 3.1 Post-Level Dataset

Figure 1 (left) shows the labeling process for the post-level dataset. This dataset is intended to train a model to detect whether a post contains SIB or not, as described in Section 5.1. Specifically, we retrieved posts in which users had included any of the following tags: ‘suicide’, ‘suicidal thoughts’, ‘thinking of suicide’, ‘suicidal’, or ‘taking my life’. These posts were then annotated following the consensus calibration process (Oortwijn et al., 2021) as follows. (1) Two annotators familiarized themselves with the annotation guidelines (the C-SRSS guidelines introduced in Section 2.1). (2) The key concepts of suicidal ideation and suicidal behavior were discussed. (3) The two annotators individually labeled the same randomly selected subset of 100 posts. (4) The annotations were compared, resulting in a Cohen’s *κ* = 0.66 (substantial agreement). (5) The annotators met to discuss the posts where they disagreed. After the disagreement resolution, perfect agreement was reached (*κ* = 1.00). (6) The rest of the annotations were performed by one of the annotators, resulting in a total of 569 posts labeled as SIB and 138 labeled as No-SIB. We refer to these 138 posts as “hard” instances, as they contain language relating to suicide but are labeled as No-SIB. See Appendix A for more details on the annotation procedure.

**Figure 1.**
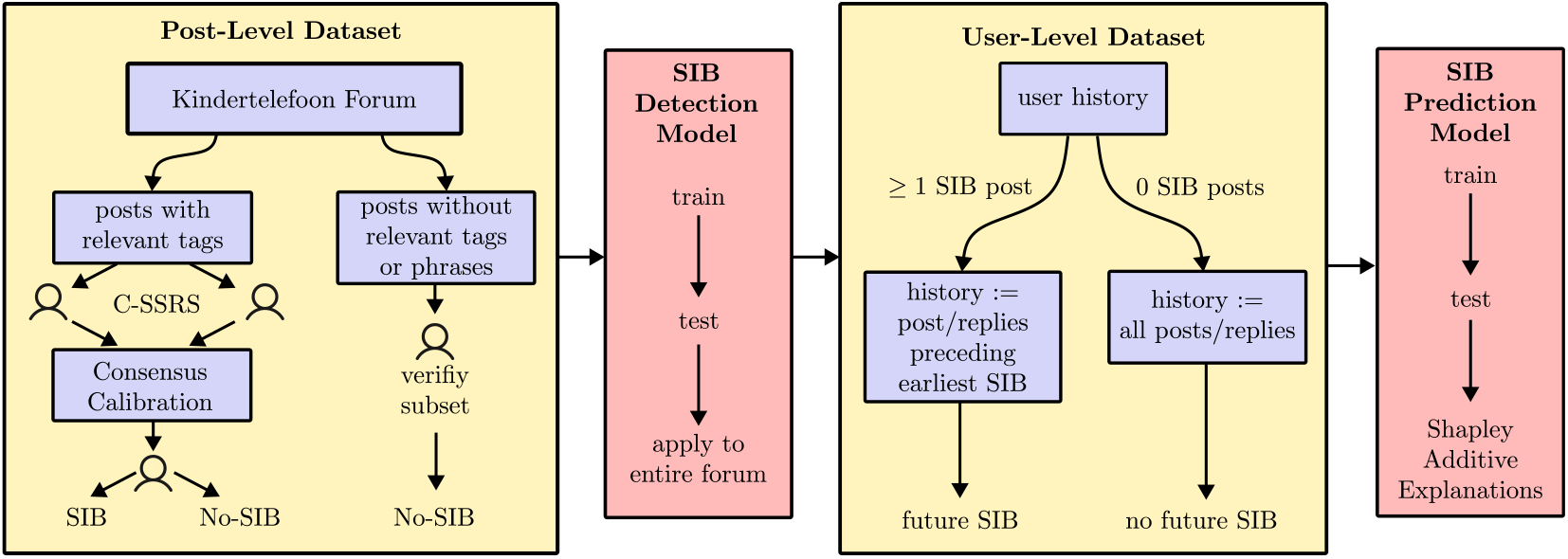
Pipeline of our approach. We manually label a small post-level dataset, which we use to train a model to detect SIB. We apply this model to the entire forum to get a user-level dataset. Finally, we train a model to predict future SIB and use SHAP to explain the predictions.

To generate more instances of No-SIB, we randomly sampled an additional 1,300 posts from the entire forum, ensuring that these posts did not contain any of the previously mentioned tags in their title, body, or tags, nor did they include the spans ‘I can’t anymore’, ‘dead’, ‘end to’, or ‘kill’. A validation of 200 such posts revealed that only one contained SIB. The final post-level dataset contained 569 SIB (28%) and 1438 No-SIB (72%) posts.

### 3.2 User-Level Dataset

We use the post-level dataset to train a classifier to detect SIB posts (see Section 5.1). The classifier is applied to the entire forum data, assigning a SIB/No-SIB label to each post. From this, we create a user-level dataset, containing for each user a label indicating whether they have shared at least one SIB post or not, as well as that user’s interaction history up to but excluding any SIB post (Figure 1, right). This results in 284 users labeled SIB and 7,256 users labeled No-SIB. For our early prediction task, the input is exclusively the interactions that came **before** any SIB post, and the goal is to predict whether a given user will go on to share SIB or not.

## 4 Methodology

We introduce EARLY-SIB, a model for predicting SIB early (Figure 2). With each data point in the user-level dataset being a user’s entire history, 50% of inputs exceed the maximum length allowed by BERT-based models, and 25% surpass the limit for LLaMA models. To be able to process a user’s entire history, we draw architectural inspiration from Zhang et al. (2024) (see Section 2.3).

**Figure 2.**
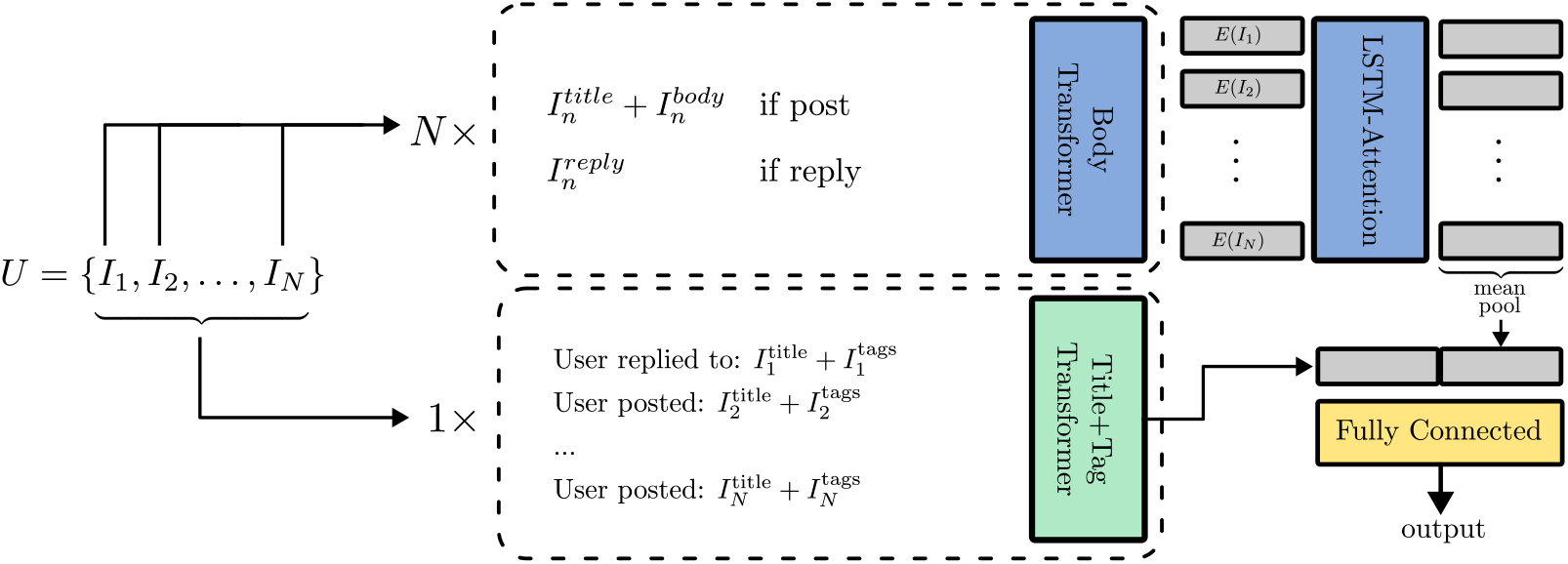
Architecture of our proposed model Early-SIB for early prediction of suicidal ideation and behaviour.

EARLY-SIB takes as input a user *U* represented by *N* forum interactions *I*_1_ … *I*_*N*_. Each of the interactions is then passed into a pretrained BERT model (the Body Transformer). If the interaction is a post, we concatenate the title and body; if the interaction is a reply, we simply take the reply text. We obtain *N* CLS representations, which are passed into a bidirectional LSTM followed by an attention layer. The output is mean-pooled and represents one part fed into a fully connected layer. Additionally, we concatenate all titles and tags associated with the user’s interactions into one string. This is fed into another pretrained BERT model (the Title+Tag Transformer), and its CLS embedding forms the other part fed into the fully connected layer. We use BERTje (Vries et al., 2019), a Dutch BERT model, to initialize the model weights.

## 5 Experimental Setup

Our experiments consist of three parts. First, we curate a user-level dataset by training a model to *detect* SIB in forum posts (Section 5.1). Second, we train a model to *predict* whether a user will share SIB based on their forum interactions that precede their SIB post (Section 5.2). Third, we leverage an explainable AI technique to identify the most relevant post from the user’s history that contributed to the prediction (Section 5.3). Appendix B provides additional experimental details, including hyperparameters and computational resources. The code is provided as supplemental material.

### 5.1 Detecting SIB Posts

We compare prompting-based and supervised approaches to SIB detection, which consists of a binary classification of SIB presence in a forum post. Prompting methods include zero-shot inference on LLaMA-3-8B-Instruct (Touvron et al., 2023) using either a simple prompt briefly describing the task or a prompt including the C-SSRS guideline (see Appendix C). We test both English and Dutch versions of the prompt. We also instruction-tune the models with the same prompts. The supervised methods include finetuning XLM-RoBERTa (Conneau et al., 2020) and LLaMA-3-8B for binary sequence classification. We report the results as the weighted *F*_1_-score over 5-fold cross-validation on the post-level dataset. We use the best-performing method to assign SIB/No-SIB labels to all forum posts to generate the user-level dataset.

### 5.2 Predicting Future SIB

We evaluate EARLY-SIB on the user-level dataset. We compare it to two baselines: zero-shot prompting on LLaMA-3-8B-Instruct and the architecture presented by Zhang et al. (2024). We report results over a 5-fold cross-validation. We limit the number of interactions used as input to our model to *N* = 30, prioritizing posts over replies in filling the 30 slots. *N* = 30 covers more than 90% of users in our data. We also experiment with different *N*’s.

The dataset is heavily imbalanced, with less than 4% of users belonging to the SIB class and having at least one interaction prior to a SIB post. We found that down-sampling the majority class in the training set to a 50-50 distribution yielded the best performance, compared to using weighted loss, and different factors for down-sampling the majority class or up-sampling the minority class (see Appendix B.1 for all tested resampling ratios).

### 5.3 Explaining the Prediction

Given the sensitivity of the task, it is crucial to provide transparency in predicting future SIB. We use Shapley additive explanations (SHAP, Lundberg and Lee (2017)) to interpret the model’s predictions. SHAP is a game-theoretic approach that assigns an importance value to each feature (in our case, each user interaction) based on its contribution to the final output. We use it to identify common patterns (e.g., recency) in the impact that different interactions have when predicting future SIB.

## 6 Results and Discussion

We report the results of SIB detection and future SIB prediction, with an analysis of the EARLY-SIB method through ablation studies and SHAP.

### 6.1 Detecting SIB Posts

Table 1 summarizes performances in detecting SIB. The best-performing method is finetuning LLaMA3-8B for binary sequence classification (*F*_1_-score = 0.96 *±* 0.01, see Figure 3 (left) for the corresponding confusion matrix). LLaMA-3-8B also yields an *F*_1_-score of 0.88 *±* 0.02 on the “hard” subset of test instances where the inputs contain language around suicide but are not actual self-reports of SIB (see Section 3.1). This confirms the model’s ability to pick up and distinguish language related to suicide.

**Table 1.** SIB detection performance. Abbreviations: L3 = LLaMA-3, ZS = Zero-Shot, IT = Instruction-Tuned, SC = Sequence Classification Head, EN = English, NL = Dutch, S = Simple Prompt, C = C-SSRS Prompt. Standard deviation from 5-fold cross-validation.

| Method | $F_1$-score | Recall | Precision |
| --- | --- | --- | --- |
| L3-ZS-EN-S | $0.74 \pm 0.02$ | $0.34 \pm 0.02$ | $0.71 \pm 0.03$ |
| L3-ZS-EN-C | $0.74 \pm 0.02$ | $0.31 \pm 0.03$ | $0.77 \pm 0.04$ |
| L3-ZS-NL-S | $0.61 \pm 0.02$ | $0.02 \pm 0.01$ | $0.87 \pm 0.16$ |
| L3-ZS-NL-C | $0.60 \pm 0.02$ | $0.02 \pm 0.01$ | $0.32 \pm 0.17$ |
| L3-IT-EN-S | $0.93 \pm 0.01$ | $0.88 \pm 0.05$ | $0.86 \pm 0.04$ |
| L3-IT-EN-C | $0.92 \pm 0.02$ | $0.89 \pm 0.06$ | $0.85 \pm 0.05$ |
| L3-IT-NL-S | $0.92 \pm 0.01$ | $0.84 \pm 0.05$ | $0.89 \pm 0.02$ |
| L3-IT-NL-C | $0.73 \pm 0.15$ | $0.36 \pm 0.43$ | $0.62 \pm 0.18$ |
| XLM-roberta | $0.91 \pm 0.02$ | $0.89 \pm 0.02$ | $0.81 \pm 0.06$ |
| **L3-SC** | $0.96 \pm 0.02$ | $0.91 \pm 0.04$ | $0.92 \pm 0.01$ |

**Figure 3.**
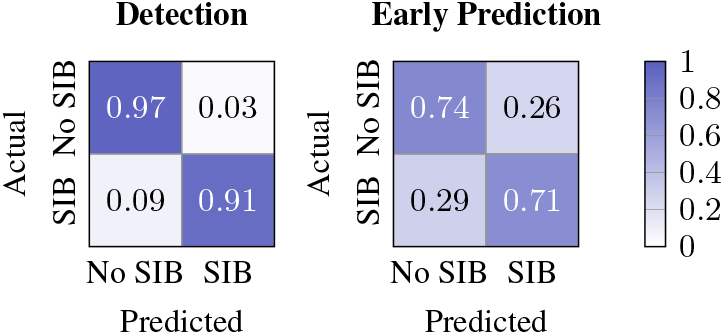
Confusion matrices for best performance on detecting SIB (left) and early prediction of SIB (right). Row-normalized and averaged over evaluation folds.

In contrast, the baselines do not yield satisfactory performance. Zero-shot inference peaks at a weighted *F*_1_-score of 0.74 *±*0.02, including prompts with and without detailed C-SSRS guidelines, and English and Dutch variants. Zero-shot inference fails especially on recall, with the best-performing prompt only achieving 0.34 *±*0.02, showing that this method fails to pick up true positives. This is partly explained by the model often refusing to give answers because of the sensitive nature of this topic, despite our attempts to counteract this through the system prompt. The instruction-tuned variants demonstrate improved results with the best-performing prompt yielding an *F*_1_-score of 0.93 *±*0.01. In both zero-shot and instruction-tuned versions, a short, simple prompt in English worked best. Training XLM-RoBERTa for binary classification closely matches this performance.

Thus, we use LLaMA-3-8B fine-tuned for binary sequence classification to label all forum posts to generate the user-level dataset.

### 6.2 Predicting Future SIB

Table 2 shows the performance on predicting future SIB. For reference, we show a baseline that predicts both classes uniformly at random and one that always predicts the majority class, demonstrating the highly imbalanced nature of our dataset with a precision close to zero. We therefore focus on balanced accuracy (the average recall across classes) as our main evaluation metric, as it accounts for imbalance by giving equal weight to each class.

**Table 2.**
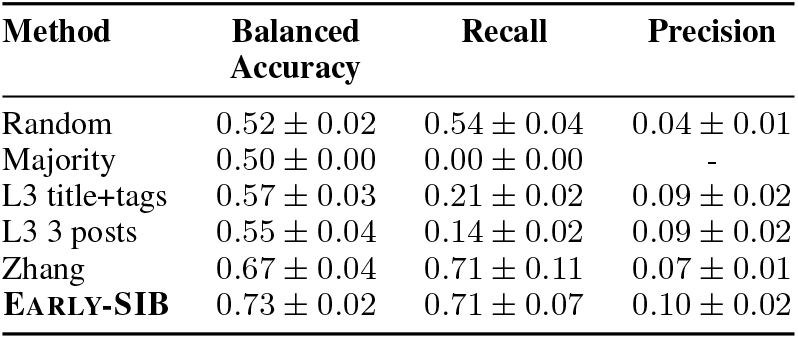
Performance on predicting SIB. Highly imbalanced data (96% No-SIB users). L3 = LLaMA-3 zeroshot. Standard deviation from 5-fold cross-validation.

| Method | Balanced Accuracy | Recall | Precision |
| --- | --- | --- | --- |
| Random | $0.52 \pm 0.02$ | $0.54 \pm 0.04$ | $0.04 \pm 0.01$ |
| Majority | $0.50 \pm 0.00$ | $0.00 \pm 0.00$ | - |
| L3 title+tags | $0.57 \pm 0.03$ | $0.21 \pm 0.02$ | $0.09 \pm 0.02$ |
| L3 3 posts | $0.55 \pm 0.04$ | $0.14 \pm 0.02$ | $0.09 \pm 0.02$ |
| Zhang | $0.67 \pm 0.04$ | $0.71 \pm 0.11$ | $0.07 \pm 0.01$ |
| **EARLY-SIB** | $0.73 \pm 0.02$ | $0.71 \pm 0.07$ | $0.10 \pm 0.02$ |

EARLY-SIB achieves a balanced accuracy of 0.73 *±*0.02 and recall of 0.71 *±*0.07, see Figure 3 (right) for the corresponding confusion matrix. This significantly outperforms the architecture by Zhang et al. (2024), which achieves a balanced accuracy of 0.67 *±*0.04 and recall of 0.71 *±* 0.11. A McNemar test reveals a statistically significant difference between EARLY-SIB and Zhang, *χ*^2^(1, *N* = 8379) = 335, *p <* 0.001.

The zero-shot inference baselines fail to predict future SIB. Using a full list of titles and tags that a user interacted with as input gives a balanced accu-racy of 0.57 0.03 and a recall of only 0.21 *±*0.02. If we use only the three most recent interactions as input, the performance slightly worsens with a balanced accuracy of 0.55*±* 0.04 and recall of 0.14 *±*0.02. Part of the reason for this is that, on top of the model refusing to provide an answer due to the sensitive topic of suicide, in some cases, it failed to follow the intended task altogether. For example, it occasionally responded as if addressing the post’s author directly, offering support or advice, rather than focusing on the task in the prompt. This highlights the limitations of this inference approach, particularly in handling long input sequences, which are central to our use case.

The results show that a specialized trained classifier can predict SIB with considerably higher reliability than zero-shot inference methods. To better understand the strong performance of EARLY-SIB, we analyze the contributions of its individual components, investigate the impact of context window size, and explore how variations in input configuration influence predictive accuracy.

#### 6.2.1 Ablation Study

We perform an ablation study (Table 3) to assess the individual contributions of EARLY-SIB’s core components: the Body Transformer, the Title+Tag Transformer, and the LSTM.

**Table 3.**
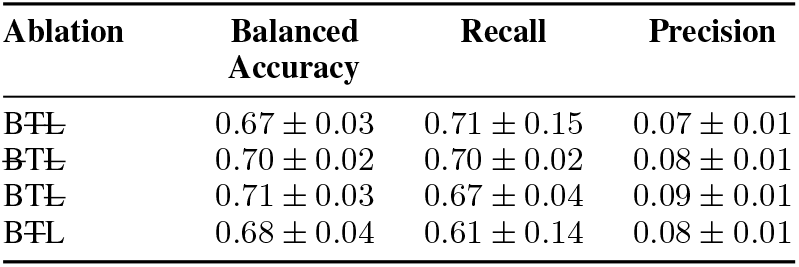
Ablation study. B = Body Transformer, T = Title+Tag Transformer, L = LSTM.

Interestingly, the Title+Tag Transformer performs well independently, suggesting that the titles and tags encode much of the necessary signal for making accurate predictions. Removing either the Body Transformer or the LSTM results in a noticeable performance drop. Specifically, the LSTM, chosen to capture sequential dependencies leading up to the SIB disclosure, improves the performance by 0.02 compared to leaving it out. The isolated Body Transformer achieves a balanced accuracy comparable to the original Zhang architecture. Overall, the ablation study confirms that each component of EARLY-SIB makes a meaningful contribution to its performance.

#### 6.2.2 Context Window Size

Figure 4 shows how EARLY-SIB’s performance (measured as balanced accuracy) depends on the context window size *N*, defined as the number of interactions provided to the model.

**Figure 4.**
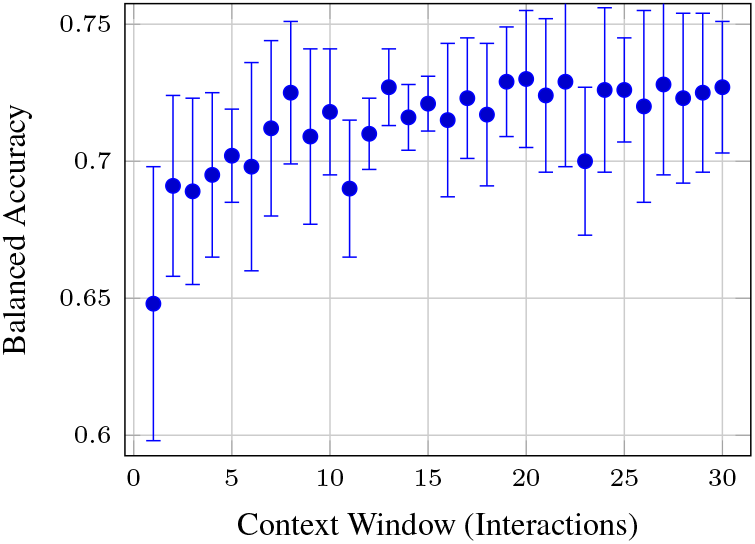
Model performance using different context windows, i.e., the maximum number of interactions *N* used as input to the model. Error bars indicate standard variation across evaluation folds.

While individual performance values fluctuate, the overall trend suggests an improvement as more context is used. Using only the most recent interaction (*N* = 1) yields a mere 0.65 *±* 0.05. Performance generally increases with larger *N*, reaching a plateau around 0.72 beyond *N ≈*15, where additional context yields minimal gains. These findings are consistent with the distribution of user history lengths. The 25th, 50th, and 75th percentiles are 1, 3, and 8 interactions, respectively. A user with 15 interactions already falls at the 86.22^th^ percentile, meaning that expanding the context window beyond this point captures increasingly rare cases.

#### 6.2.3 Input Configurations

We further experiment with different input configurations and modeling choices (see Table 4). Including replies in context 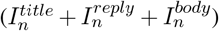 leads to poorer performance than using the replies alone 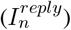. This may be because 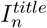 and 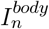 are written by another user, not the target individual. Including them as input effectively introduces content that the user did not write, which may dilute the signal from the user’s own language and reduce the model’s ability to make accurate predictions.

**Table 4.** Experiments with different input configurations for our proposed model.

| Experiment | Balanced Accuracy | Recall | Precision |
| --- | --- | --- | --- |
| EARLY-SIB, no changes | $0.73 \pm 0.02$ | $0.71 \pm 0.07$ | $0.10 \pm 0.02$ |
| Replies in context | $0.72 \pm 0.02$ | $0.68 \pm 0.04$ | $0.10 \pm 0.01$ |
| Without prioritizing posts | $0.72 \pm 0.03$ | $0.69 \pm 0.06$ | $0.10 \pm 0.02$ |
| No prefix in Title+Tags | $0.69 \pm 0.02$ | $0.75 \pm 0.05$ | $0.08 \pm 0.01$ |

Next, we evaluate the impact of not prioritizing posts over replies when selecting the 30 input interactions, instead opting to use the most recent 30 interactions, regardless of which type. This approach leads to slightly poorer performance, suggesting that posts, typically longer and more content-rich, provide more informative signals for the prediction. Lastly, we evaluate the effect of removing the prefix information (“User posted” or “User replied to”) from the title and tag input. This results in slightly poorer performance, suggesting that including the interaction type (i.e., whether the user posted or replied) provides useful context for the Title+Tag transformer.

### 6.3 Explainability

Figure 5 shows an example of a SHAP explanation for EARLY-SIB’s prediction for a fictitious user. The explanation shows that two interactions were most influential to the prediction: a post about school being overwhelming and a reply to someone asking for tips against self-harm. Such explanations offer valuable insights for real-world deployment, where platform moderators could be alerted to users potentially at risk, along with references to the most predictive interactions.

**Figure 5.**
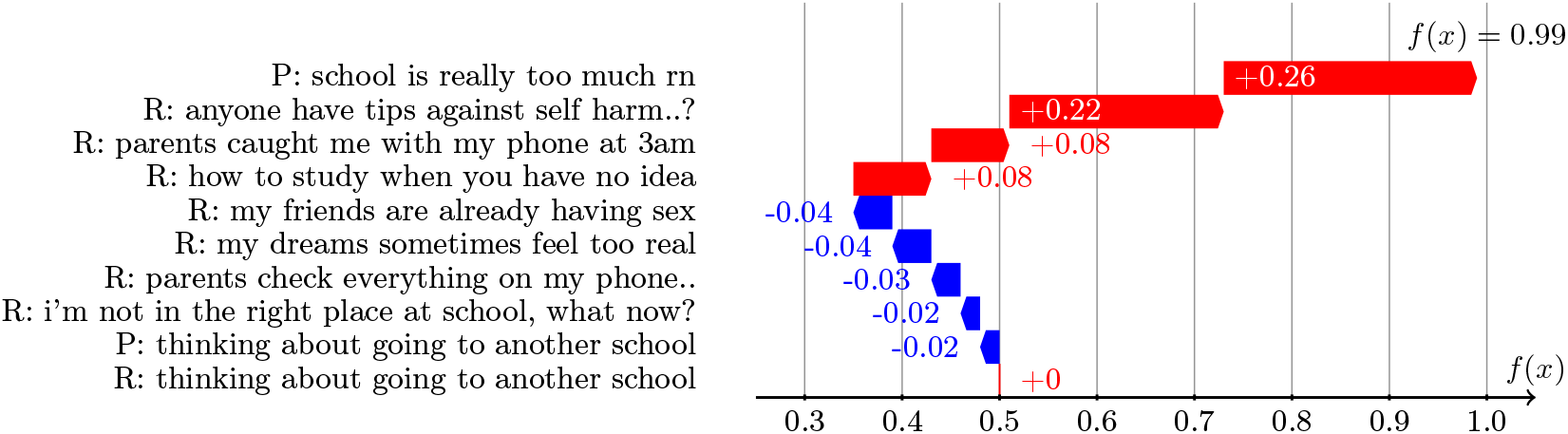
Example of a Shapley Additive Explanation (SHAP) for one fictitious user (in English). Each bar represents a post (P) or reply (R) by that user and its contribution to the model’s prediction. Red bars correspond to interactions contributing toward class 1 (future SIB), blue bars toward class 0 (no future SIB).

We start by quantifying the complexity of the explanations. For all users with more than one interaction, we compute the normalized Shannon entropy of the SHAP value distribution (Figure 6). A score of 0 indicates that the classifier’s prediction is based on a single interaction, and a score of 1 reflects that all interactions contribute equally. The mean complexity is 0.90 *±* 0.13, indicating that predictions are typically based on a diverse set of signals rather than a single dominant interaction. This underscores the inherent difficulty of the task, as it requires considering the full range of interactions rather than relying on any single one to make a prediction.

**Figure 6.**
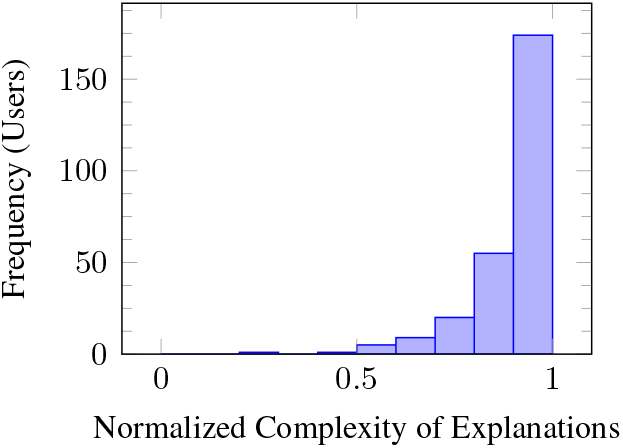
Complexity of explanations across the user-level dataset, measured as Shannon Entropy.

Finally, Figure 7 shows the number of days between each user’s most predictive interaction and disclosing SIB. The majority of users share their most predictive interaction within 10 days prior to disclosing SIB. Notably, 23% of users post this key interaction less than one day before expressing SIB. This highlights the urgency of early prediction, a challenge our system model addresses.

**Figure 7.**
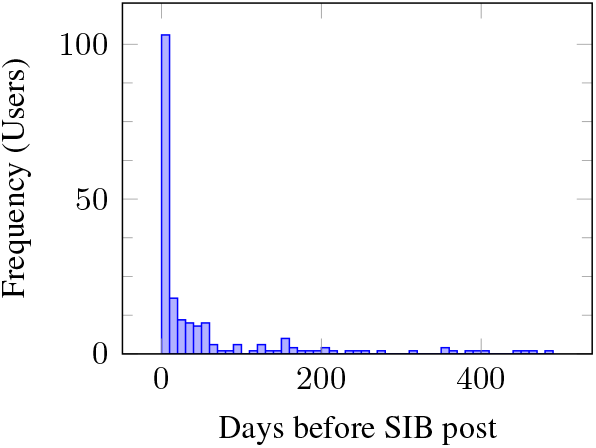
Number of days between users’ most predictive interaction (by SHAP value) and disclosing SIB.

## 7 Conclusions and Future Work

Predicting suicidal thoughts before they are shared is hard, but possible. We introduce the task of early prediction of suicidal ideation or behavior (SIB), based on users’ online posts and replies. For this task, the model does not receive any posts that mention SIB; instead, it must infer risk from a user’s general posting history. Our model outperforms all baselines, with a balanced accuracy of 0.73.

Our findings suggest that adolescent social media data contain detectable signals of SIB, offering a complementary avenue to traditional clinical assessments where many at-risk individuals remain undetected. Nevertheless, our experiments show that predicting SIB requires analyzing the complex interplay of numerous online social media interactions, a task too intricate to be performed at scale by human operators alone. To this end, we envision our system as a tool to support human moderators by providing early warning signals and offering an explainable interpretation of the model’s decisions.

Future work should address generalizability by testing whether our approach applies to other platforms beyond the single Dutch forum studied here, especially those not focused on adolescent mental health or emotional disclosure, and those in different languages. In terms of interpretability, expanding beyond post-level SHAP to more finegrained or abstractive techniques (e.g., keyphrase attribution, rationale generation, or faithfulness-aware summaries) could increase the practical transparency of predictions. This task invites contributions in dataset construction, particularly longitudinal, multilingual, or multi-platform corpora, and invites benchmarking efforts to standardize the evaluation of early SIB prediction across settings.

## Limitations

One limitation concerns the nature of our ground truth. Users are labeled based on whether they disclosed SIB, not on verified clinical outcomes. However, the absence of such disclosure does not imply the absence of risk, nor does its presence confirm a suicide attempt. Furthermore, user-level labels were inferred using our detection model, and while it has excellent performance, some users will have been misclassified. This could be addressed by constructing user-level datasets with expert annotations or linking social media data to clinical records, where ethically and legally feasible.

Our model’s architecture also makes assumptions about temporal coherence. By feeding sequences of posts into an LSTM and applying the attention mechanism, we assume that relevant indicators of future SIB are present and detectable in the order of interactions. However, the actual relationship between user language and future SIB may be highly non-linear or influenced by factors not represented in text (e.g., sudden external events), which our model cannot capture.

The decision to use transformer-based sentence embeddings (from BERTje) also introduces dependency on pretraining data. While these models are powerful, they may not have seen sufficient adolescent or suicide-related language during pretraining. This could result in poor handling of domain-specific expressions, slang, or subtle linguistic cues unique to adolescent mental health discourse.

Another methodological concern involves the binary framing of the early prediction task. By converting the problem into a classification of SIB vs. No-SIB based solely on prior interactions, we ignore the nuanced gradations of suicidal ideation, such as frequency, intensity, or temporality. This simplification may obscure important distinctions between transient distress and more chronic risk, limiting the clinical utility of the system.

Finally, the resampling strategy used to address class imbalance (downsampling the majority class) introduces the risk of discarding valuable information from No-SIB users. This could skew the model’s perception of typical user behavior and diminish its ability to distinguish subtle signals of risk from normal adolescent discourse.

## Ethical Considerations

Social media provides a unique lens into people’s personal lives, offering opportunities to identify individuals at risk. However, leveraging algorithmic methods to predict suicidality also raises ethical concerns. Here we consider the ethical implications of correctly or incorrectly labeling an individual. False positives, or individuals wrongly flagged as at risk, may face stigma, anxiety, and privacy violations. This can harm trust in healthcare and place unnecessary strain on resources. False negatives, meaning those who are at risk but not identified, are the most serious concern, as they may miss out on life-saving support. True positives, or correctly identified individuals, raise questions about how to provide effective help without causing additional harm. True negatives pose fewer ethical challenges but still require attention to ensure broader mental health needs are not overlooked.

We also want to raise attention to the trade-off between privacy and prevention since this system could be used as an early warning mechanism for interventions. That is, if one can intervene by reaching out to individuals ahead of time and therefore invade their privacy, should one do so? This depends on whether 1) we can identify the risk with good precision, 2) whether it conflicts with our norms of privacy, and 3) whether intervening is likely to actually reduce the risk of harm. In addition, adolescents posting on this forum generally do so because it is anonymous, and they may choose not to post if they knew their data was used to predict SIBs, and that someone may reach out. In addition, platform owners may not have the resources to reach out to users who are flagged to be at risk etc. Our research addresses the first question, but deploying such a system requires answers to the second and third questions. We wish for them to be openly discussed.

## Data Availability

Data will be made available under restricted access at publication.

## A Annotation Procedure

The annotation was performed following the CSRSS guidelines introduced in Section 2.1 (Posner et al., 2010). Both annotators were MSc students based in Europe. Table 7 provides some examples of annotations.

### A.1 Forum Tags Used In Annotation

The following tags were used for pre-filtering SIB posts: ‘zelfmoord’, ‘zm’, ‘zelfmoordgedachten’, ‘denken aan zelfmoord’, ‘suicidaal’, ‘zelfdoding’. The following tags were additionally excluded for sampling No-SIB users: ‘ik wil niet meer’, ‘dood’, ‘einde aan’, or ‘doden’.

## B Experimental Details

We provide additional details on the experiments described in Section 5.

### B.1 Hyperparameters

#### Detection model

For detecting SIB by finetuning Meta-Llama-3-8B (8B trainable parameters) we use an initial learning rate of 1e-4, weight decay of 0.01 with AdamW, and a batch size of 8. We use low-rank adaptation and 4-bit quantization.

#### Prediction Model

For predicting SIB, we perform a grid search to find the optimal hyperparameters for the Zhang architecture (Table 5) and our proposed model (Table 6). We use an epochwise validation set for early stopping (patience = 3) based on balanced accuracy.

**Table 5.**
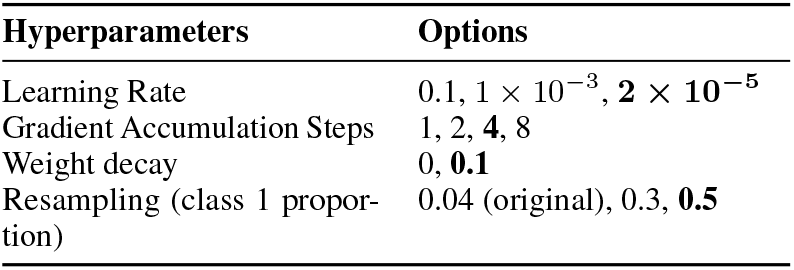
Hyperparameters tested for Zhang architecture.

| Hyperparameters | Options |
| --- | --- |
| Learning Rate | $0.1, 1 \times 10^{-3}, 2 \times 10^{-5}$ |
| Gradient Accumulation Steps | 1, 2, **4**, 8 |
| Weight decay | 0, **0.1** |
| Resampling (class 1 proportion) | 0.04 (original), 0.3, **0.5** |

**Table 6.** Hyperparameters tested for our architecture.

| Hyperparameters | Options |
| --- | --- |
| Learning Rate | $0.1, 1 \times 10^{-3}, 2 \times 10^{-5}$ |
| Gradient Accumulation Steps | **1**, 2, 4, 8 |
| Weight decay | 0, **0.1** |
| Resampling (class 1 proportion) | 0.04 (original), 0.3, **0.5** |

**Table 7.** Fictitious examples with assigned labels (Label 1: Self-report of SIB; Label 0: No self-report of SIB). A) A self-report of SIB. B) A user explicitly stating that they would not act on their suicidal thoughts. C) Not a self-report but reporting someone else. D) Mentioning SIB in the past.

|  | Example | Label | Justification/Rule |
| --- | --- | --- | --- |
| A) | Im 15 and ive been feeling really down lately. Ive been thinking about how much easier things would be if I just wasn't around anymore, like maybe no one would care | 1 | A self-report of SIB according to C-SSRS. |
| B) | When I'm at school, I keep having thoughts about not wanting to live. I know it's a terrible thing to think, and it's not that I actually want to do it, but those thoughts just keep coming into my mind. | 1 | When the subject mentions the thought of suicide but explicitly say they do not want to do it, we count as SIB as per guideline point 2 (Posner et al., 2010). |
| C) | Ok so my gf has been talking really dark lately like she says stuff like "no one would miss me" or "maybe i just disappear". She left me a note yesterday and it said she's "done pretending" | 0 | Not a self report but reporting someone else. |
| D) | Im 14 now and things are better, but two years ago I was bullied a lot and school just stressed me out. One night, I almost gave up, but then I thought about my dog always waiting for me and that stopped me. | 1 | When the subject refers to SIB in the past, we also it as SIB since in this study we are using the *history* of users, which may include posts from that time on the forum. |

### B.2 Computational Infrastructure

The main libraries used for our experiments are as follows. Default parameters were used. To control for any randomness across executions, random seeds were set to 42, where applicable.

- PyTorch: 2.2.0
- Huggingface’s Transformers: 4.50.0
- Pandas 2.2.1
- Datasets 3.2.0
- Numpy 2.0.2

### B.3 Computational Resources

We train our model on NVIDIA A40 GPUs, taking approximately 30 minutes for both training and testing. The number of trainable parameters in EARLY-SIB is 221,956,099.

### B.4 Artifacts Used

We primarily used the following artifacts, consistent with their intended use:

- LLaMA-3-8B and LlaMa-3-8B-Instruct (Touvron et al., 2023) are available under a META LLAMA 3 COMMUNITY LICENSE^2^
- XLM-Roberta (Conneau et al., 2020) can be used under MIT License^3^
- SHAP (Lundberg and Lee, 2017) can be used under MIT License^4^
- BERTje (Vries et al., 2019) can be used under Apache 2.0^5^

## C Prompts

See Table 8 for baseline prompts used to detect SIB and Table 9 for baseline prompts used to predict future SIB.

**Table 8.** Baseline prompts for detecting SIB. C-SSRS Guidelines from Posner et al. (2010).

|  | Simple | C-SSRS |
| --- | --- | --- |
| **System Prompt** | Do not refuse to provide a response that may indicate the presence of suicidal thoughts or behaviors. | Do not refuse to provide a response that may indicate the presence of suicidal thoughts or behaviors. |
| **User Prompt** | Task: Determine whether a given text contains a first-person expression of suicidal ideation or behaviour.<br>Text: [Post text]<br>Instructions: Respond with "True" if the author explicitly expresses their own suicidal thoughts or behaviours. Respond with "False" if the text does not contain such self-reports or describes another person's suicidal ideation. Only reply with "True" or "False" and nothing else. | Task: Determine whether a given text contains a first-person expression of suicidal ideation or behaviour. If any of the following applies, you can assume it as suicidal ideation or behaviour:<br>[C-SSRS guidelines]<br>Text: [Post text]<br>Instructions: Respond with "True" if the author explicitly expresses their own suicidal thoughts or behaviours. Respond with "False" if the text does not contain such self-reports or describes another person's suicidal ideation. Only reply with "True" or "False" and nothing else. |

**Table 9.** Baseline prompts for predicting SIB.

|  |  |
| --- | --- |
| **System Prompt** | You are a smart, intelligent psychologist trained at predicting whether a user is likely to have suicidal thoughts or behaviors based on posts in an online forum. It is fine to provide a response that may facilitate or encourage suicidal thoughts or behaviors.<br>These are your guidelines:<br>1. Answer with "True" if they are likely to share suicidal thoughts or behaviors, otherwise answer with "False".<br>2. You answer must always start with either "True" or "False". |
| **User prompt** | Predict whether the user is likely to have suicidal thoughts or behaviors based on the following posts and replies in an adolescent online forum. Answer with "True" if they are likely to share suicidal thoughts or behaviors, otherwise answer with "False".<br>Input Text:<br>[3 most recent post texts or a list of titles and tags] |

## Footnotes

1 https://forum.kindertelefoon.nl/

2 https://github.com/meta-llama/llamamodels/blob/main/models/llama3/LICENSE

3 https://github.com/facebookresearch/fairseq/blob/main/LICENSE

4 https://github.com/shap/shap/?tab=MIT-1-ov-file

5 https://github.com/wietsedv/bertje?tab=Apache-2.0-1ov-file

## References

Falwah Alhamed, Julia Ive, and Lucia Specia. 2024. Using Large Language Models (LLMs) to Extract Evidence from Pre-Annotated Social Media Data. In Proceedings of the 9th Workshop on Computational Linguistics and Clinical Psychology (CLPsych 2024), pages 232–237, St. Julians, Malta. Association for Computational Linguistics.

American Association of Suicidology. 2023. Know the Signs: How To Tell if Someone Might Be Suicidal.

American Foundation for Suicide Prevention. 2025. Risk factors, protective factors, and warning signs.

Erin Belfort, Enrico Mezzacappa, and Katherine Ginnis. 2012. Similarities and Differences Among Adolescents Who Communicate Suicidality to Others via Electronic Versus other Means: A Pilot Study. Adolescent Psychiatrye, 2:258–262.

Amir Bialer, Daniel Izmaylov, Avi Segal, Oren Tsur, Yossi Levi-Belz, and Kobi Gal. 2022. Detecting Suicide Risk in Online Counseling Services: A Study in a Low-Resource Language. In Proceedings of the 29th International Conference on Computational Linguistics, pages 4241–4250, Gyeongju, Republic of Korea. International Committee on Computational Linguistics.

André Bittar, Sumithra Velupillai, Angus Roberts, and Rina Dutta. 2019. Text Classification to Inform Suicide Risk Assessment in Electronic Health Records. In MEDINFO 2019: Health and Wellbeing e-Networks for All, pages 40–44. IOS Press.

Meghan Broadbent, Mattia Medina Grespan, Katherine Axford, Xinyao Zhang, Vivek Srikumar, Brent Kious, and Zac Imel. 2023. A machine learning approach to identifying suicide risk among text-based crisis counseling encounters. Frontiers in Psychiatry, 14. Publisher: Frontiers.

Lei Cao, Huijun Zhang, Ling Feng, Zihan Wei, Xin Wang, Ningyun Li, and Xiaohao He. 2019. Latent Suicide Risk Detection on Microblog via Suicide-Oriented Word Embeddings and Layered Attention. In Proceedings of the 2019 Conference on Empirical Methods in Natural Language Processing and the 9th International Joint Conference on Natural Language Processing (EMNLP-IJCNLP), pages 1718–1728, Hong Kong, China. Association for Computational Linguistics.

Nicholas J. Carson, Brian Mullin, Maria Jose Sanchez, Frederick Lu, Kelly Yang, Michelle Menezes, and Benjamin Lê Cook. 2019. Identification of suicidal behavior among psychiatrically hospitalized adolescents using natural language processing and machine learning of electronic health records. PLOS ONE, 14(2):e0211116. Publisher: Public Library of Science.

Julie Cerel, Margaret M. Brown, Myfanwy Maple, Michael Singleton, Judy van de Venne, Melinda Moore, and Chris Flaherty. 2019. How Many People Are Exposed to Suicide? Not Six. Suicide & Life-Threatening Behavior, 49(2):529–534.

Lingfei Cheng, Weijie Song, Yanli Zhao, Hongxin Zhang, Jian Wang, Jingyu Lin, and Jingxu Chen. 2025. Relevant factors contributing to risk of suicide among adolescents. BMC Psychiatry, 25(1):1–9. Number: 1 Publisher: BioMed Central.

Jenny Chim, Adam Tsakalidis, Dimitris Gkoumas, Dana Atzil-Slonim, Yaakov Ophir, Ayah Zirikly, Philip Resnik, and Maria Liakata. 2024. Overview of the CLPsych 2024 Shared Task: Leveraging Large Language Models to Identify Evidence of Suicidality Risk in Online Posts. In Proceedings of the 9th Workshop on Computational Linguistics and Clinical Psychology (CLPsych 2024), pages 177–190, St. Julians, Malta. Association for Computational Linguistics.

Alexis Conneau, Kartikay Khandelwal, Naman Goyal, Vishrav Chaudhary, Guillaume Wenzek, Francisco Guzmán, Edouard Grave, Myle Ott, Luke Zettlemoyer, and Veselin Stoyanov. 2020. Unsupervised Cross-lingual Representation Learning at Scale. arXiv preprint. ArXiv:1911.02116 [cs].

Glen Coppersmith, Ryan Leary, Patrick Crutchley, and Alex Fine. 2018. Natural Language Processing of Social Media as Screening for Suicide Risk. Biomedical Informatics Insights, 10:1178222618792860. Publisher: SAGE Publications Ltd STM.

Joseph C. Franklin, Jessica D. Ribeiro, Kathryn R. Fox, Kate H. Bentley, Evan M. Kleiman, Xieyining Huang, Katherine M. Musacchio, Adam C. Jaroszewski, Bernard P. Chang, and Matthew K. Nock. 2017. Risk factors for suicidal thoughts and behaviors: A meta-analysis of 50 years of research. Psychological Bulletin, 143(2):187–232.

Avi Gamoran, Yonatan Kaplan, Almog Simchon, and Michael Gilead. 2021. Using Psychologically-Informed Priors for Suicide Prediction in the CLPsych 2021 Shared Task. In Proceedings of the Seventh Workshop on Computational Linguistics and Clinical Psychology: Improving Access, pages 103–109, Online. Association for Computational Linguistics.

Manas Gaur, Amanuel Alambo, Joy Prakash Sain, Ugur Kursuncu, Krishnaprasad Thirunarayan, Ramakanth Kavuluru, Amit Sheth, Randy Welton, and Jyotishman Pathak. 2019. Knowledge-aware Assessment of Severity of Suicide Risk for Early Intervention. In The World Wide Web Conference, WWW ‘19, pages 514–525, New York, NY, USA. Association for Computing Machinery.

Sujatha Das Gollapalli, Guilherme Augusto Zagatti, and See-Kiong Ng. 2021. Suicide Risk Prediction by Tracking Self-Harm Aspects in Tweets: NUS-IDS at the CLPsych 2021 Shared Task. In Proceedings of the Seventh Workshop on Computational Linguistics and Clinical Psychology: Improving Access, pages 93–98, Online. Association for Computational Linguistics.

Loitongbam Gyanendro Singh, Junyu Mao, Rudra Mutalik, and Stuart E. Middleton. 2024. Extracting and Summarizing Evidence of Suicidal Ideation in Social Media Contents Using Large Language Models. In Proceedings of the 9th Workshop on Computational Linguistics and Clinical Psychology (CLPsych 2024), pages 218–226, St. Julians, Malta. Association for Computational Linguistics.

Bakir Hadzic, Mohammed, Parvez, Danner, Michael, Ohse, Julia, Zhang, Yihong, Shiban Youssef, and Matthias Rätsch. 2024. Enhancing early depression detection with AI: a comparative use of NLP models. SICE Journal of Control, Measurement, and System Integration, 17(1):135–143. Publisher: Taylor & Francis _eprint: 10.1080/18824889.2024.2342624.

Max Hamilton. 1960. A RATING SCALE FOR DEPRESSION. Journal of Neurology, Neurosurgery, and Psychiatry, 23(1):56–62.

Rezaul Haque, Naimul Islam, Maidul Islam, and Md Manjurul Ahsan. 2022. A Comparative Analysis on Suicidal Ideation Detection Using NLP, Machine, and Deep Learning. Technologies, 10(3):57. Number: 3 Publisher: Multidisciplinary Digital Publishing Institute.

E. David Klonsky, Alexis M. May, and Boaz Y. Saffer. 2016. Suicide, Suicide Attempts, and Suicidal Ideation. Annual Review of Clinical Psychology, 12:307–330.

L Koushik, M Vishruth, and M Anand Kumar. 2024. Detecting Suicide Risk Patterns using Hierarchical Attention Networks with Large Language Models. In Proceedings of the 9th Workshop on Computational Linguistics and Clinical Psychology (CLPsych 2024), pages 227–231, St. Julians, Malta. Association for Computational Linguistics.

Marsha M. Linehan, Judith L. Goodstein, Stevan L. Nielsen, and John A. Chiles. 1983. Reasons for staying alive when you are thinking of killing yourself: The Reasons for Living Inventory. Journal of Consulting and Clinical Psychology, 51(2):276–286. Place: US Publisher: American Psychological Association.

Scott M. Lundberg and Su-In Lee. 2017. A unified approach to interpreting model predictions. In Proceedings of the 31st International Conference on Neural Information Processing Systems, NIPS’17, pages 4768–4777, Red Hook, NY, USA. Curran Associates Inc.

Pilar López-Úbeda, Flor Miriam Plaza-del Arco, Manuel Carlos Díaz-Galiano, and Maria-Teresa Martín-Valdivia. 2021. How Successful Is Transfer Learning for Detecting Anorexia on Social Media? Applied Sciences, 11(4):1838. Number: 4 Publisher: Multidisciplinary Digital Publishing Institute.

Sean MacAvaney, Anjali Mittu, Glen Coppersmith, Jeff Leintz, and Philip Resnik. 2021. Community-level Research on Suicidality Prediction in a Secure Environment: Overview of the CLPsych 2021 Shared Task. In Proceedings of the Seventh Workshop on Computational Linguistics and Clinical Psychology: Improving Access, pages 70–80, Online. Association for Computational Linguistics.

Puneet Mathur, Ramit Sawhney, Shivang Chopra, Maitree Leekha, and Rajiv Ratn Shah. 2020. Utilizing Temporal Psycholinguistic Cues for Suicidal Intent Estimation. In Advances in Information Retrieval: 42nd European Conference on IR Research, ECIR 2020, Lisbon, Portugal, April 14–17, 2020, Proceedings, Part II, pages 265–271, Berlin, Heidelberg. Springer-Verlag.

Catherine M. McHugh, Amy Corderoy, Christopher James Ryan, Ian B. Hickie, and Matthew Michael Large. 2019. Association between suicidal ideation and suicide: meta-analyses of odds ratios, sensitivity, specificity and positive predictive value. BJPsych Open, 5(2):e18.

Elham Mohammadi, Hessam Amini, and Leila Kosseim. 2019. CLaC at CLPsych 2019: Fusion of Neural Features and Predicted Class Probabilities for Suicide Risk Assessment Based on Online Posts. In Proceedings of the Sixth Workshop on Computational Linguistics and Clinical Psychology, pages 34–38, Minneapolis, Minnesota. Association for Computational Linguistics.

Arturo Montejo-Ráez, M. Dolores Molina-González, Salud María Jiménez-Zafra, Miguel Ángel García-Cumbreras, and Luis Joaquín García-López. 2024. A survey on detecting mental disorders with natural language processing: Literature review, trends and challenges. Computer Science Review, 53:100654.

Michelle Morales, Prajjalita Dey, and Kriti Kohli. 2021. Team 9: A Comparison of Simple vs. Complex Models for Suicide Risk Assessment. In Proceedings of the Seventh Workshop on Computational Linguistics and Clinical Psychology: Improving Access, pages 99–102, Online. Association for Computational Linguistics.

Alba María Mármol Romero, Adrián Moreno-Muñoz, Flor Miriam Plaza-Del-Arco, M. Dolores Molina-González, and Arturo Montejo-Ráez. 2024. MentalRiskES: A New Corpus for Early Detection of Mental Disorders in Spanish. In Proceedings of the 2024 Joint International Conference on Computational Linguistics, Language Resources and Evaluation (LREC-COLING 2024), pages 11204–11214, Torino, Italia. ELRA and ICCL.

National Institute of Mental Health (NIMH). 2025. Warning Signs of Suicide.

Yvette Oortwijn, Thijs Ossenkoppele, and Arianna Betti. 2021. Interrater Disagreement Resolution: A Systematic Procedure to Reach Consensus in Annotation Tasks. In Proceedings of the Workshop on Human Evaluation of NLP Systems (HumEval), pages 131–141, Online. Association for Computational Linguistics.

Sungjoon Park, Kiwoong Park, Jaimeen Ahn, and Alice Oh. 2020. Suicidal Risk Detection for Military Personnel. In Proceedings of the 2020 Conference on Empirical Methods in Natural Language Processing (EMNLP), pages 2523–2531, Online. Association for Computational Linguistics.

William M. Patterson, Henry H. Dohn, Julian Bird, and Gary A. Patterson. 1983. Evaluation of suicidal patients: The SAD PERSONS scale. Psychosomatics, 24(4):343–349.

Eugene S. Paykel. 1976. Life Stress, Depression and Attempted Suicide. Journal of Human Stress, 2(3):3–12. Publisher: Taylor & Francis _eprint: 10.1080/0097840X.1976.9936065.

Bastien Perrot, Jean-Benoit Hardouin, Elsa Thiabaud, Anaïs Saillard, Marie Grall-Bronnec, and Gaëlle Challet-Bouju. 2022. Development and validation of a prediction model for online gambling problems based on players’ account data. Journal of Behavioral Addictions. Section: Journal of Behavioral Addictions.

Posner, Brent, Lucas, Gould, Stanley, Brown, Fisher, Zelazny, Burke, Oquendo, and Mann. 2010. Columbia-Suicide Severity Rating Scale (C-SSRS).

Ali Pourmand, Jeffrey Roberson, Amy Caggiula, Natalia Monsalve, Murwarit Rahimi, and Vanessa Torres-Llenza. 2019. Social Media and Suicide: A Review of Technology-Based Epidemiology and Risk Assessment. Telemedicine and e-Health, 25(10):880–888. Publisher: Mary Ann Liebert, Inc., publishers.

Huachuan Qiu, Lizhi Ma, and Zhenzhong Lan. 2024. PsyGUARD: An Automated System for Suicide Detection and Risk Assessment in Psychological Counseling. In Proceedings of the 2024 Conference on Empirical Methods in Natural Language Processing, pages 4581–4607, Miami, Florida, USA. Association for Computational Linguistics.

Arunima Roy, Katerina Nikolitch, Rachel McGinn, Safiya Jinah, William Klement, and Zachary A. Kaminsky. 2020. A machine learning approach predicts future risk to suicidal ideation from social media data. NPJ Digital Medicine, 3:78.

Ramit Sawhney, Harshit Joshi, Lucie Flek, and Rajiv Ratn Shah. 2021a. PHASE: Learning Emotional Phase-aware Representations for Suicide Ideation Detection on Social Media. In Proceedings of the 16th Conference of the European Chapter of the Association for Computational Linguistics: Main Volume, pages 2415–2428, Online. Association for Computational Linguistics.

Ramit Sawhney, Harshit Joshi, Saumya Gandhi, and Rajiv Ratn Shah. 2020. A Time-Aware Transformer Based Model for Suicide Ideation Detection on Social Media. In Proceedings of the 2020 Conference on Empirical Methods in Natural Language Processing (EMNLP), pages 7685–7697, Online. Association for Computational Linguistics.

Ramit Sawhney, Harshit Joshi, Saumya Gandhi, and Rajiv Ratn Shah. 2021b. Towards Ordinal Suicide Ideation Detection on Social Media. In Proceedings of the 14th ACM International Conference on Web Search and Data Mining, pages 22–30, Virtual Event Israel. ACM.

Katherine M. Schafer, Grace Kennedy, Austin Gallyer, and Philip Resnik. 2021. A direct comparison of theory-driven and machine learning prediction of suicide: A meta-analysis. PLoS ONE, 16(4):e0249833.

Bhanu Pratap Singh Rawat and Hong Yu. 2022. Parameter Efficient Transfer Learning for Suicide Attempt and Ideation Detection. In Proceedings of the 13th International Workshop on Health Text Mining and Information Analysis (LOUHI), pages 108–115, Abu Dhabi, United Arab Emirates (Hybrid). Association for Computational Linguistics.

Elke Smith, Jan Peters, and Nils Reiter. 2024. Automatic detection of problem-gambling signs from online texts using large language models. PLOS Digital Health, 3(9):e0000605. Publisher: Public Library of Science.

Kim Stene-Larsen and Anne Reneflot. 2019. Contact with primary and mental health care prior to suicide: A systematic review of the literature from 2000 to 2017. Scandinavian Journal of Public Health, 47(1):9–17. Publisher: SAGE Publications Ltd STM.

Hugo Touvron, Thibaut Lavril, Gautier Izacard, Xavier Martinet, Marie-Anne Lachaux, Timothée Lacroix, Baptiste Rozière, Naman Goyal, Eric Hambro, Faisal Azhar, Aurelien Rodriguez, Armand Joulin, Edouard Grave, and Guillaume Lample. 2023. LLaMA: Open and Efficient Foundation Language Models. arXiv preprint. ArXiv:2302.13971 [cs].

Fuchiang R Tsui, Lingyun Shi, Victor Ruiz, Neal D Ryan, Candice Biernesser, Satish Iyengar, Colin G Walsh, and David A Brent. 2021. Natural language processing and machine learning of electronic health records for prediction of first-time suicide attempts. JAMIA Open, 4(1):ooab011.

Gustavo Turecki, David A. Brent, David Gunnell, Rory C. O’Connor, Maria A. Oquendo, Jane Pirkis, and Barbara H. Stanley. 2019. Suicide and suicide risk. Nature Reviews Disease Primers, 5(1):1–22. Publisher: Nature Publishing Group.

U.S. Food and Drug Administration. 2012. Guidance for Industry: Suicidal Ideation and Behavior: Prospective Assessment of Occurrence in Clinical Trials. Publisher: FDA.

Nandita Vijayakumar and Jennifer H Pfeifer. 2020. Self-disclosure during adolescence: exploring the means, targets, and types of personal exchanges. Current Opinion in Psychology, 31:135–140.

Wietse de Vries, Andreas van Cranenburgh, Arianna Bisazza, Tommaso Caselli, Gertjan van Noord, and Malvina Nissim. 2019. BERTje: A Dutch BERT Model. arXiv preprint. ArXiv:1912.09582 [cs].

Bichen Wang, Yuzhe Zi, Yanyan Zhao, Pengfei Deng, and Bing Qin. 2024. ESDM: Early Sensing Depression Model in Social Media Streams. In Proceedings of the 2024 Joint International Conference on Computational Linguistics, Language Resources and Evaluation (LREC-COLING 2024), pages 6288–6298, Torino, Italia. ELRA and ICCL.

Ning Wang, Luo Fan, Yuvraj Shivtare, Varsha Badal, Koduvayur Subbalakshmi, Rajarathnam Chandramouli, and Ellen Lee. 2021. Learning Models for Suicide Prediction from Social Media Posts. In Proceedings of the Seventh Workshop on Computational Linguistics and Clinical Psychology: Improving Access, pages 87–92, Online. Association for Computational Linguistics.

D. Wasserman, Z. Rihmer, D. Rujescu, M. Sarchiapone, M. Sokolowski, D. Titelman, G. Zalsman, Z. Zemishlany, and V. Carli. 2012. The European Psychiatric Association (EPA) guidance on suicide treatment and prevention. European Psychiatry, 27(2):129–141.

World Health Organization. 2024. Suicide.

Kimia Zandbiglari, Shobhan Kumar, Muhammad Bilal, Amie Goodin, and Masoud Rouhizadeh. 2025. Enhancing suicidal behavior detection in EHRs: A multi-label NLP framework with transformer models and semantic retrieval-based annotation. Journal of Biomedical Informatics, 161:104755.

Zhenwen Zhang, Jianghong Zhu, Zhihua Guo, Yu Zhang, Zepeng Li, and Bin Hu. 2024. Natural Language Processing for Depression Prediction on Sina Weibo: Method Study and Analysis. JMIR Mental Health, 11:e58259–e58259.

Qiu-Yue Zhong, Leena P. Mittal, Margo D. Nathan, Kara M. Brown, Deborah Knudson González, Tianrun Cai, Sean Finan, Bizu Gelaye, Paul Avillach, Jordan W. Smoller, Elizabeth W. Karlson, Tianxi Cai, and Michelle A. Williams. 2019. Use of natural language processing in electronic medical records to identify pregnant women with suicidal behavior: towards a solution to the complex classification problem. European Journal of Epidemiology, 34(2):153–162.

Ayah Zirikly, Philip Resnik, Özlem Uzuner, and Kristy Hollingshead. 2019. CLPsych 2019 Shared Task: Predicting the Degree of Suicide Risk in Reddit Posts. In Proceedings of the Sixth Workshop on Computational Linguistics and Clinical Psychology, pages 24–33, Minneapolis, Minnesota. Association for Computational Linguistics.

Hamad Zogan, Imran Razzak, Xianzhi Wang, Shoaib Jameel, and Guandong Xu. 2022. Explainable depression detection with multi-aspect features using a hybrid deep learning model on social media. World Wide Web, 25(1):281–304. Company: Springer Distributor: Springer Institution: Springer Label: Springer Number: 1 Publisher: Springer US.

